# PRESCRIBING PRACTICES AND CLINICAL IMPACT OF NEXT GENERATION SEQUENCING IN ROUTINE PRACTICE IN SOLID TUMORS – REAL WORLD EXPERIENCE IN LMIC

**DOI:** 10.1101/2024.07.31.24311267

**Authors:** Adeeba Zaki, Nida E Zehra, Aqsa Sohail, Zeeshan Ansar, Munira Moosajee

## Abstract

Molecular characterization of disease is essential for precision medicine due to novel predictive biomarkers. Multiple next-generation sequencing (NGS) platforms are available, but their expense and clinical utility vary. Even if a targetable mutation is detected, corresponding drugs may not be available or affordable. No prior studies in Pakistan have focused on integrating NGS results into patient care to assist with therapeutic decision-making and survival outcomes.

This retrospective study aimed to evaluate the molecular profiling and therapeutic implications of NGS testing across solid tumors. It included all patients with histologically proven malignancy (metastatic or non-metastatic) who had NGS analysis at Aga Khan University Hospital (AKUH) from June 1, 2020, to June 1, 2023. Foundation One was the NGS platform used. From 2020 to 2023, 192 patients underwent NGS. The majority were male (55.2%) and aged over 50 years (71.9%). The most common indications for NGS were carcinoma of unknown primary (CUP) and lung cancers, representing 26% and 25% respectively, followed by colon (9%) and breast cancers (8%). Most patients had metastatic disease (98.4%). Common mutations in lung cancer were EGFR (16.3%) and KRAS G12C (14.3%). In unknown primary, breast, and colon cancers, the most common mutations were BRAF (8%), PIK3CA (18%), and KRAS (42.1%), respectively. Microsatellite instability (MSI) testing was performed in 95% of patients, with 6% being MSI high. Actionable alterations were detected in 31.8% of patients, but only 17.2% received genotype-matched treatment, mostly as a first-line treatment for lung cancer. The primary barriers were drug availability and affordability.

Our results show that the implementation of NGS analysis supports clinical decision making. However, these results were applicable to a small percentage of patients. For better compliance and applicability, drug availability and cost of treatment needs to be addressed

## Introduction

Cancer is a heterogenous disease. Prescence of subpopulation of tumour cells with distinct genotype and phenotype leads to divergent biological behavior within a primary tumor and its metastases, or between tumors of the same histopathological subtype [1]. Genetic variations in both the cancer and inherited genomes are informative for hereditary cancer risk, prognosis, and treatment strategies. Given the increasing number of potential targets for cancer therapies, novel applications have been developed that provide information about the tumour genome to guide therapeutic selection. The application of genetic information to assist with the clinical management of patients with cancer is rapidly expanding into routine care [2].

There are numerous genetic testing platforms that can guide cancer care from single gene testing by PCR (polymerase chain reaction) or FISH (Fluorescence in situ hybridization) to comprehensive next generation sequencing. Next generation sequencing (NGS) allows rapid simultaneous sequencing of DNA or RNA samples with a higher sensitivity and provides prognostic information on sensitivity or resistance to drugs [3]. Sequencing applications, both genome-wide and targeted, are revealing complex mutational signatures associated with different types of cancers that are now driving both research and therapeutic decisions [4]. Studies have evaluated the decision impact of molecular profiling on treatment and their effect on overall survival [5].

Studies have evaluated the cost and impact of NGS use at national level on treatment modification and survival [6]. Jones et al. report a statistically significant difference in survival for patients who showed actionable alterations with subsequent targeted therapy in comparison with patients who, despite showing actionable alterations, did not receive targeted therapy and patients who did not show any actionable alterations on NGS testing in gynecological malignancies [7]. In advanced breast cancer clinical application of NGS, real world data revealed that only 4.7% of patients received molecular guided therapy [8].

In resource limited countries, routine use of NGS testing is not possible due to cost and availability of testing facility and subsequent drugs. Therefore, we felt that it was essential to evaluate the prescribing practices of NGS in solid malignancies with a focus on the clinical impact of NGS results in patient care to assist with therapeutic decision making. To our knowledge this is the first study evaluating molecular profiling and its therapeutic implication across solid tumors in Pakistan

## Material and Methods

### Patients and sample

For this retrospective observational study, all the patients with histologically proven malignancy in whom tumor genomic profiling test ordered from June 2020 till June 2023 were included. There were no limitations for tumor type, treatment line, metastatic vs non-metastatic disease, performance status or organ function. Patients were tested with either Foundation One CDx test, in house targeted hotspot panel (limited gene testing) via PCR or FISH and/ or Microsatellite instability (MSI) testing by PCR. Foundation One CDx, sequences genes from solid tumor tissue sample and reporting the detected alteration (mutation, amplification, deletion, fusion and also tumor mutational burden) along with suggested targeted therapies against each detected alteration.

For this retrospective study, the need for individual patient consent was waived by the ethics committee due to the use of de-identified data. All data were anonymized to ensure confidentiality and privacy. As the study involved the analysis of previously collected data, the ethics committee waived the requirement for obtaining new written or verbal consent from participants. When the genomic profiling tests were initially ordered, patients provided informed written consent to have data from their medical records used in research. The study protocol, including the waiver of consent, was reviewed and approved by the ethics committee of The Aga Khan University and considered under exempted study, ensuring that all ethical considerations were addressed.

### Statistical Analysis

Data was analyzed using STATA 16.0. Mean and Standard Deviation or median and intra quartile range (IQR) were calculated for quantitative continuous variables based on the normality of the variables. Frequency and percentages were calculated for categorical variables. Confounders were controlled through stratification; post stratification Chi-square test was applied. Cox proportional hazard regression was used for multivariable analysis to determine the influence of the following variables: matched and nonmatched treatment given before and after NGS analysis, line of treatment at NGS analysis on survival for the total population. A p-value of less than and equal to 0.05 was considered as statistically significant.

## Results

367 patients underwent genomic profiling from June 2020 to June 2023 at the Aga Khan University Hospital. Clinical and demographic characteristics of patients.

192 patients underwent genomic (Next generation sequencing) testing via Foundation One CDx panel (FMI). 13 had limited targeted hotspot testing and 162 had MSI testing alone. Majority were male (58.8%) while 41% were female. On age stratification, FMI testing was done mostly in patients who were > 50 years while MSI alone was done in patients between 30-50 years presenting predominantly with non-metastatic malignancy. 22 malignancies were found in which NGS and MSI was advised by treating physician.

The most common indication was carcinoma of unknown primary (CUP) and lung cancer representing 26 % and 25% respectively. This was followed by colon cancer (9%), breast cancer (8%).MSI testing was done solely as an only hotspot test in around 44% of patients, mostly those with colon cancer. Inhouse targeted hotspot test was offered in 13 patients, mostly done in patients with colon cancer. Majority of patients had metastatic disease. NGS testing was recommended in first line of treatment after the diagnosis in majority of the patients. Actionable alterations were detected in 31.8% of which only 17.2% patients received genotype-matched treatment. This was most commonly done in first line treatment for lung cancer. Table 1

**Table 1:** Demographics.

|  | **NGS -FMI**<br>**(N=192) %** | **MSI- AKU**<br>**(N=162) %** | **NGS -AKU**<br>**(N=13) %** |
| --- | --- | --- | --- |
| **Age (Yrs.)** |  |  |  |
| <30 | 9 (4.7%) | 10 (6.2%) | 0 |
| 30-50 | 45 (23.4%) | 103 (63.6%) | 5 (38.5%) |
| >50 | 138 (71.9%) | 49 (30.2%) | 8 (61.5%) |
| **Sex** |  |  |  |
| Female | 86 (44.8%) | 60 (37.0%) | 5 (38.5%) |
| Male | 106 (55.2%) | 102 (63.0%) | 8 (61.5%) |
| **Primary malignancy** |  |  |  |
| Bone cancer | 2 (1.0%) | - |  |
| Breast cancer | 16 (8.3%) | - | 1 (7.7%) |
| Cervical cancer | 3 (1.6%) | - |  |
| Colon cancer | 19 (9.9%) | 80 (49.4%) | 10 (76.9%) |
| Esophageal cancer | 3 (1.6%) | 7 (4.3%) |  |
| Gall Bladder cancer | 4 (2.1%) | 5 (3.1%) |  |
| gastric cancer | 1 (0.5%) | 12 (7.4%) |  |
| Head and Neck cancer | 2 (1.0%) | - |  |
| Kidney Cancer | 1 (0.5%) | - |  |
| Lung cancer | 49 (25.5%) | - |  |
| Melanoma | 1 (0.5%) | - | 1 (7.7%) |
| Ovarian cancer | 4 (2.1%) | 2 (1.2%) |  |
| Pancreatic cancer | 13 (6.8%) | 7 (4.3%) | 1 (7.7%) |
| Prostate cancer | 4 (2.1%) | 5 (3.1%) |  |
| Rectal cancer | 4 (2.1%) | 36 (22.2%) |  |
| Skin cancer | 1 (0.5%) | - |  |
| Soft Tissue sarcoma | 5 (2.6%) | - |  |
| Thyroid cancer | 3 (1.6%) | - |  |
| Unknown primary | 50 (26.0%) | - |  |
| Urothelial cancer | 1 (0.5%) | - |  |
| Uterine cancer | 6 (3.1%) | 7 (4.3%) |  |
| Neuroendocrine Tumor | - | 1 (0.6%) |  |
| **Stage** |  |  |  |
| Metastatic | 189 (98.4%) | 104 (64.2%) | 13 (100%) |
| Non-metastatic | 3 (1.6%) | 58 (35.8%) | 0 |
| **Line of treatment (At the time of NGS)** |  |  |  |
| First | 187 (97.4%) | 153 (94.4%) | 13 (100%) |
| Second | 5 (2.6%) | 9 (5.6%) | 0 |
| **MSI testing** | 183 (95.3%) |  |  |
| **MSI-H** | 6 (3.2%) | 27 (16.7%) | - |
| **MSS** | 177 (96.7%) | 135 (83.3%) | - |
| **Tumour mutational burden** |  |  |  |
| <10 Muts/Mb | 123 (64.1%) | - | - |
| >10 Muts/Mb | 23 (12.0%) | - | - |
| 0 Muts/Mb | 46 (24.0%) | - | - |
| **Actional alteration** | 61 (31.8%) | - | 3 (23.07%) |
| **No Alteration (Wild type)** | 131 (68.2%) | - | 10 (76.9%) |
| **Genotype matched treatment** |  |  |  |
| N/A | 3 (1.6%) | - |  |
| No | 156 (81.3%) | 113 (69.7%) | 12 (92.3) |
| Yes | 33 (17.2%) | 49 (30.2%) | 1 (7.6%) |
| **Line of treatment (first/second/third or beyond)** |  |  |  |
| First Line | 17 (8.9%) | 35 (21.6%) | 1 (7.6%) |
| N/A | 157 (81.8%) | - | - |
| Second line | 18 (9.4%) | 14 (8.6%) | - |

### GENOMIC LANDSCAPE

#### MSI testing

MSI testing was done as a part of Foundation one CDx panel and also as a targeted hotspot test via PCR at institute. Most common indication was colon cancer (49%) followed by rectal cancer (22%) and gastric cancer (7%). In colon cancer, 28.7% patients are found to have MSI -H while 71.2% are MSS and most commonly performed in stage II colon cancer. In rectal cancer majority of the patients were MSS (92%) and mostly done in metastatic disease. Matched treatment according to instability testing was given in all patients with colon cancer. MSI testing in patients other than colon cancer was done in metastatic presentation. Table 2

**Table 2:** MSI testing MSI (N=162)

| Primary malignancy | MSI testing<br>No. (%) | MSI-H<br>No. (%) | MSS<br>No. (%) | Matched treatment<br>No. (%) |
| --- | --- | --- | --- | --- |
| Colon cancer | 80<br>(49.4%) | 23 (28.7%) | 57 (71.2%) | 46 (93.9%) |
| Esophageal cancer | 7 (4.3%) | 1 (14.2%) | 6 (85.7%) | 2 (4.1%) |
| Gall Bladder cancer | 5 (3.1%) | 5 (100%) | 0 (0%) | 0 (0%) |
| Gastric cancer | 12 (7.4%) | 1 (8.3%) | 11 (91.6%) | 1 (2.0%) |
| Ovarian cancer | 2 (1.2%) | 0 (0%) | 2 (100%) | 0 (0%) |
| Pancreatic cancer | 7 (4.3%) | 0 (0%) | 7 (100%) | 0 (0%) |
| Prostate cancer | 5 (3.1%) | 0 (0%) | 5 (100%) | 0 (0%) |
| Rectal cancer | 36<br>(22.2%) | 1 (2.7%) | 35 (97.2%) | 0 (0%) |
| Uterine cancer | 7 (4.3%) | 1 (14.2%) | 6 (85.7%) | 0 (0%) |
| Neuroendocrine tumor | 1 (0.6%) | 0 (0%) | 1 (100%) | 0 (0%) |

**Table 3:** Cancer and Alteration.

| Cancer and Alteration | % |
| --- | --- |
| **Pancreatic Cancer** |  |
| CDK4- amplification | 8% |
| ERBB2- amplification | 15% |
| KRAS- G12D | 46% |
| MDM2- amplification | 8% |
| NF1- rearrangement | 8% |
| STK11- loss | 8% |
| **Cervical Cancer** |  |
| MET- amplification/ ARID1A- S11fs*91 | 33% |
| PIK3CA | 33% |
| PIK3CA- | 33% |
| **Esophageal Cancer** |  |
| ARID1A- Y551fs*68 | 33% |
| CCND1- amplification | 33% |
| ERBB2- amplification/ MYC- amplification | 33% |
| **Gall Bladder Cancer** |  |
| (PD-L1)- amplification/ (PD-L2)-<br>amplification/ KRAS- amplification, | 25% |
| EGFR- amplification-equivocal/ NF1-<br>F2156L | 25% |
| ERBB2-amplification- | 50% |
| **Ovarian Cancer** |  |
| RAD51B- R47* | 25% |
| CBL- C381Y | 25% |
| NF1- L190*/ MYC- amplification-<br>equivocal/ TP53- R280* | 25% |
| **Prostate Cancer** |  |
| APC- F1954fs*16/ PIK3R1- T576del,<br>rearrangement intron 7/ PTEN- loss | 25% |
| AR- amplification/ CTNNB1- S45P-<br>subclonal/ PTEN- loss/ RET- amplification | 25% |
| MYC- amplification/ TMPRSS2-<br>TMPRSS2-ERG fusion | 25% |
| NF1- rearrangement intron 13 | 25% |
| **Rectal Cancer** |  |
| KRAS- G12C | 25% |
| NRAS- Q61K | 25% |
| RNF43- R221fs*39 | 25% |
| **Soft tissue sarcoma** |  |
| CDK4- amplification/ MDM2- amplification/ | 60% |
| EWSR1- EWSR1-ATF1 fusion | 20% |
| PTEN- loss exons 3-9 | 20% |
| **Thyroid cancer** |  |
| BRCA1- S1253fs*10/ AKT2- amplification/ AKT3- amplification-equivocal/ KEAP1- Y300*/ MTAP- loss exons 5-8 | 33% |
| NRAS- Q61R | 33% |
| RET- M918T/ NF1- W221* | 33% |
| **Uterine cancer** |  |
| PIK3CA | 50% |
| FBXW7- R479*/ PIK3CA- | 17% |
| PIK3R1 | 17% |

#### Cancer types and molecular targets and implication on treatment

22 different cancers genomic profiling were studied. Most common were the Lung cancer, carcinoma of unknown primary, breast and colon cancer. Each with different demographic details, genomic alteration (either targetable or non-targetable), line of treatment and matched treatment as per result. Most actionable alteration were detected in lung cancer. Mostly the patients had upfront testing when diagnosed with metastatic disease.

49 patients with lung cancer, 50 with carcinoma of unknown primary, 16 with breast cancer, 92 patients with colon cancer underwent NGS (Foundation one) testing. In all cancer type mostly patients were male expect those with breast cancer with age greater than 50 years. Most common mutation detected in lung cancer was EGFR Exon 19 deletion (16.3%), KRAS-G12C amplification (14.3%), ERBB amplification (8.2%) followed by EGFR L858R, CDK 4 amplification, BRAF. (Graph 1). Common mutation in carcinoma of unknown primary were BRAF/ MYC amplification (8%), CDK-4 amplification (8%) followed by EGFR amplification, NF-1 loss (6%). (Graph 2). In breast cancer, common mutation was PIK3CA amplification, PTEN (18%) followed by BRCA 2 (12.5%) and BRCA 1 (6.3%) (Graph 3). And in colon cancer common mutation detected were KRAS and TP53 (42.1%) followed by TP53 alone (26.3%). All the patients were Microsatellite stable (MSS) with Tumor mutational burden (TMB) < 10 mut/Mb. Only 10.5% of the patients were MSI-H in colon cancer. (Graph 4).

Actionable mutation was detected in 30%, 16%, 37% and 47% in lung, carcinoma of unknown primary, breast cancer and colon cancer respectively. Genotype matched treatment was received in 30%, 6%, 12.5% and 36% respectively. Most likely limitation for not receiving genotype matched treatment was due to non – availability of drug and non-affordability.

**Graph 1:**
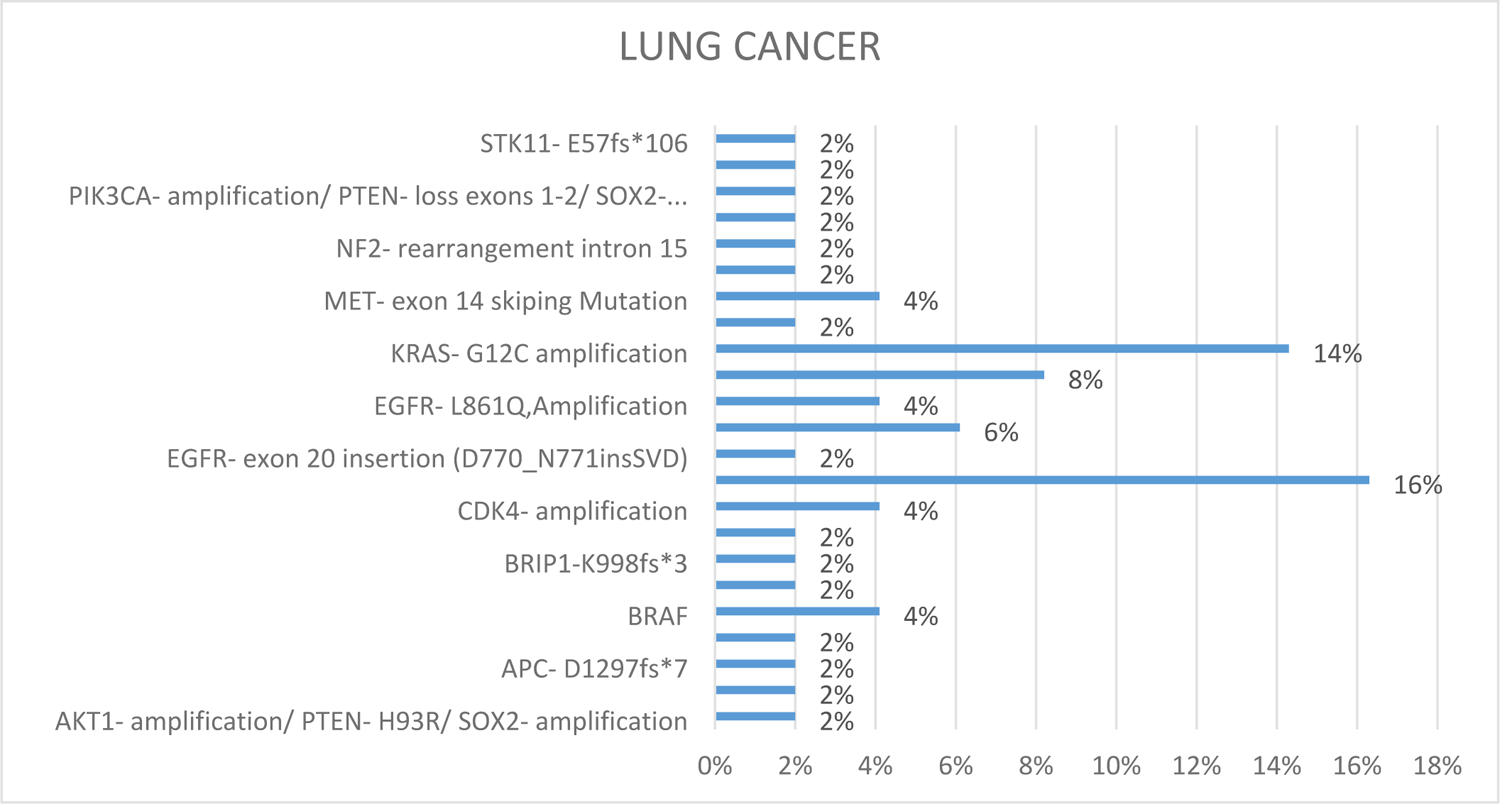
Molecular Alteration detected in NGS – Lung cancer

**Graph 2:**
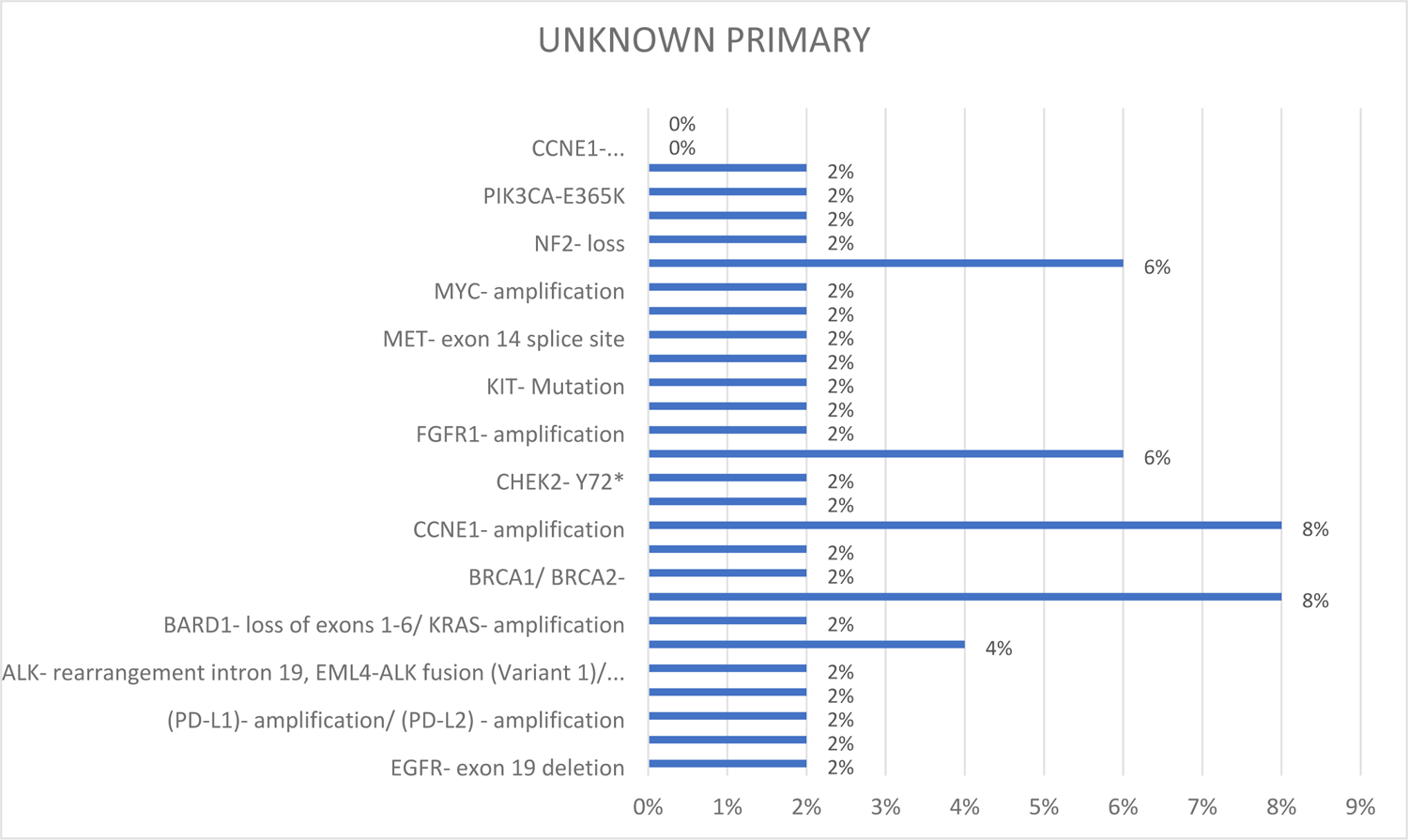
Molecular Alteration detected in NGS-Carcinoma of Unknown Primary.

**Graph 3:**
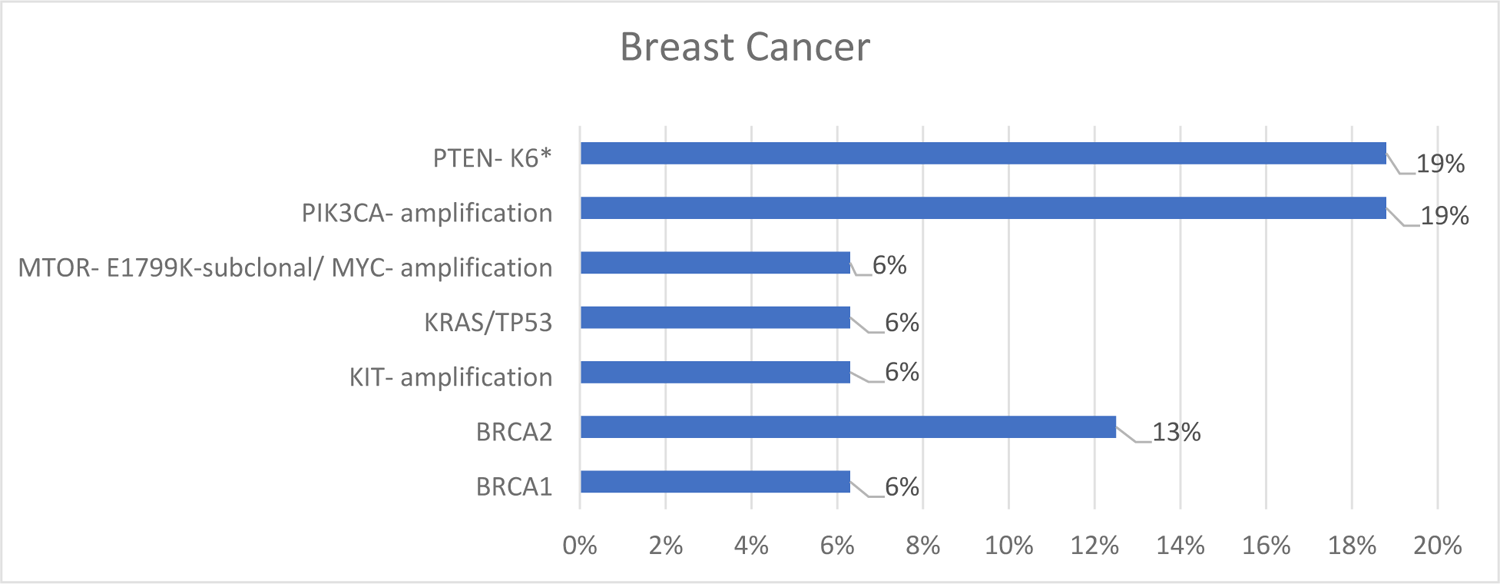
Molecular Alteration detected in NGS-Breast Cancer

**Graph 4:**
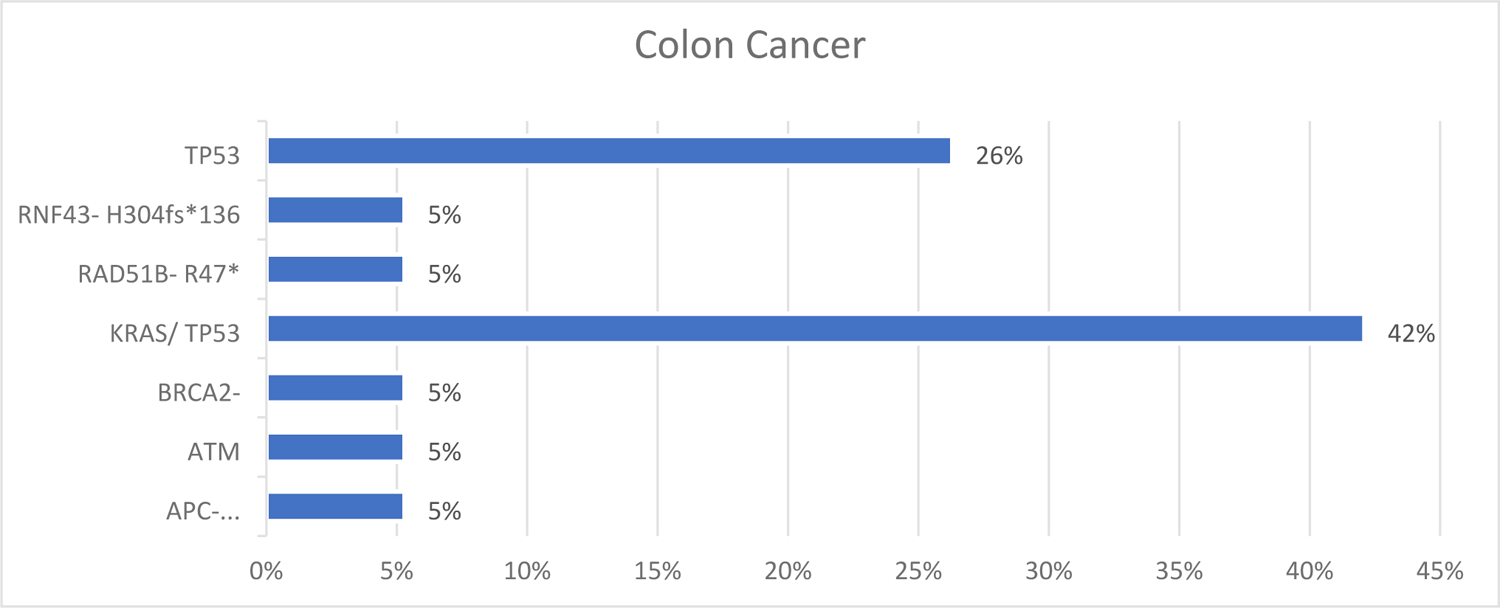
Molecular Alteration detected in NGS-Breast Cancer

#### Other cancer types

17 other different cancers genomic profiling were studied. No actionable mutation was detected in majority of cancer. While in other cancers like esophageal cancer (3 patients), Gall bladder cancer (4 patients), rectal cancer (4 patients), skin cancer (1 patient), thyroid cancer (3 patients) and uterine cancer (6 patients), in which actionable alteration were detected but no genotype matched treatment was given mainly due to non-availability of drugs. Table-3

## Discussion

The success in drug interaction associated with genomic alteration have led to implementation of NGS testing with the capability to sequence many genes for specific cancer in a short period of time. In real world medical practice NGS panel testing ordered by oncologist increasingly but still there is still controversy that whether molecular profiling is needed upfront in all the oncological patients, this issue is still unresoved [9]. In 2020 NGS recommendation were published which proposed that NGS testing must be focued on advanced cancer in which several molecular markers are relavent for the initial treatment like colon cancer, lung cancer, malenoma, unknown primary tumors [10]. Our current study have appraised the integration of NGS into clinical perspective of patient care which can help in assessing therapeutic decision making among solid tumors.

Carcinoma of unknown primary always remains a unique challenge to the clinician as represented as a heterogenous group of malignancies with distinct disease course and biology. Studies have shown that progress in genomic profiling elaborate further insight into downstream pathways and cellular system which further aid in subclassification into tumor origin that may sometime help as a therapeutic target [11]. retrospective study on CUP revealed that most common actionable alteration identified were PIK3CA, BRCA2, KRAS mutation, ERBB2 amplification and high TMB. Median overall survival for those who treated with molecular guided therapy was 23.6 months as compared to historical data using conventional therapy [12]. In our study only 16% of patients with carcinoma of unknown primary found to have actionable alteration and of which 6% received molecular based treatment. Common mutations were BRAF/ MYC amplification, CDK-4 amplification, EGFR amplification, NF-1 loss.

In lung cancer (specifically stage IV Non-small cell lung cancer (NSCLC), molecular testing upfront before starting treatment has become standard of care. NGS though saves time and is cost-effective when we compared with gene testing done sequentially but in our resource constraint setting, we are obliged to get sequential testing in some patients. Presley et al. reported 29% of actionable mutation via NGS testing [13] also report 61% targeted therapy usage [14]. Our study revealed 55% of actionable target alteration which is similar to reported but only 30% of patients received genotype matched treatment which is either due to unavailability of drug or non-affordability. Furthermore, our study did not compare survival outcome between patients undergoing NGS testing versus sequential molecular testing.

From the discovery of molecular biomarkers KRAS/NRAS in colon cancer with confers resistance to anti EGFR therapy we have moved to different molecular subtype of metastatic colorectal cancer with different treatment options. Therefore, molecular profiling became essential, and the guideline recommends that all the patients with metastatic colorectal cancer must be tested for. Our study also revealed that the most common mutation detected was KRAS/TP53 and 10% MSI-H with is similar to reported in the studies [15].

Improved outcomes were shown in cancer particularly lung cancer, clinical trial screening or rare cancer for making therapeutic decision association with NGS testing when it was performed initially. Median PFS was found to be almost a year in cancer patients with a good performance status and those with early in course of metastatic assessment as compared to those with rapidly progressing tumors and with poor performance status.

NGS testing impact actually has some limitation that it only focuses on gene alteration but has not taken into account the patient’s general health and on the outcomes associated with test and also improvement in survival has not been addressed clearly. Our study although limited in sample size offers an overview of in which cancer subtype NGS is mostly ordered by oncologist. Our observation strengthens the need of NGS in advanced cancer and the need for improving the understanding of genomic drug association.

Our results show that the implementation of NGS analysis does support clinical decision making. However, the results are applicable in small percentage of patients. For better compliance and applicability, drug availability and cost of treatment need to be addressed.

## Data Availability

N/A

